# Sequencing SARS-CoV-2 Genomes from Saliva

**DOI:** 10.1101/2021.06.21.21259289

**Authors:** Tara Alpert, Chantal B.F. Vogels, Mallery I. Breban, Mary E. Petrone, Yale IMPACT Research Team, Anne L. Wyllie, Nathan D. Grubaugh, Joseph R. Fauver

## Abstract

Genomic sequencing is crucial to understanding the epidemiology and evolution of SARS-CoV-2. Often, genomic studies rely on remnant diagnostic material, typically nasopharyngeal swabs, as input into whole genome SARS-CoV-2 next-generation sequencing pipelines. Saliva has proven to be a safe and stable specimen for the detection of SARS-CoV-2 RNA via traditional diagnostic assays, however saliva is not commonly used for SARS-CoV-2 sequencing. Using the ARTIC Network amplicon-generation approach with sequencing on the Oxford Nanopore MinION, we demonstrate that sequencing SARS-CoV-2 from saliva produces genomes comparable to those from nasopharyngeal swabs, and that RNA extraction is necessary to generate complete genomes from saliva. In this study, we show that saliva is a useful specimen type for genomic studies of SARS-CoV-2.

## Introduction

Genomic studies of SARS-CoV-2 are critical to the collective understanding and control of the COVID-19 pandemic. Unbiased genomic sequencing identified a novel betacoronavirus (SARS-CoV-2) in the bronchoalveolar lavage fluid of an individual with pneumonia in December, 2019^1^. This first SARS-CoV-2 genome sequence paved the way for the design of numerous vaccines^2^, the development of diagnostic assays^3^, and targeted sequencing approaches^4^. Since then, more than 2 million SARS-CoV-2 consensus sequence genomes have been uploaded and shared on GISAID (gisaid.org) as of June 21, 2021. Open data sharing has facilitated wide scale viral surveillance that has led to the identification of multiple variants of concern and shaped both clinical and public health approaches to the treatment and control of COVID-19^5^.

Workflows for whole-genome SARS-CoV-2 sequencing begin with sample collection, typically from discarded clinical diagnostic specimens. While nasopharyngeal (NP) swabs are still considered as the ‘gold standard’ for SARS-CoV-2 diagnostic testing, we have shown that saliva is a sensitive^6,7^ and stable^8^ sample type, which can be reliably self-collected^9^, for the detection of SARS-CoV-2 RNA by RT-qPCR diagnostic assays in the absence of RNA extraction^10^. As saliva is increasingly being used in diagnostic testing programs, we sought to determine if saliva samples can be used to generate high-quality SARS-CoV-2 genomes.

In this study, we consider two common measures of genome quality: depth of coverage (i.e. the number of reads aligning to the genome) and breadth of coverage (i.e. genome completeness). Intrinsic viral diversity, polymerase errors, and sequencing artifacts introduce heterogeneity into sequencing data, making it difficult to determine the consensus nucleotide identity with few reads. The ARTIC Network protocol only assigns a nucleotide identity to positions with at least 20x coverage (i.e. 20 or more reads align to a given position in the genome) to increase confidence in the consensus genome sequence. The completeness of a sequenced SARS-CoV-2 genome often depends on the viral load and sample quality. Because SARS-CoV-2 RNA typically makes up a small proportion of RNA in clinical diagnostic specimens, PCR amplification increases the amount of genomic material available for sequencing. However, due to the highly multiplexed nature of the ARTIC Network PCR approach, where more than a hundred individual primer sets are pooled in a single reaction, a slight imbalance in priming efficiency can lead to unequal read distribution and result in incomplete genomes. Thus, genome completeness is rarely 100%, rather genomes that are 80-99% complete are considered high-quality for the purposes of this study.

We compared SARS-CoV-2 genome quality from i) saliva and NP swabs of varying viral concentrations from a hospital cohort, ii) paired saliva and NP swab samples from the same individual, and iii) saliva samples with and without RNA extraction. Our results show that saliva performs similarly to NP swabs using both random and paired samples, genome coverage is strongly correlated with viral load and data quantity, and that performing RNA extraction from saliva drastically improves genome completeness. These data demonstrate that high-quality SARS-CoV-2 genomes can be readily sequenced from saliva samples.

## Results

### SARS-CoV-2 genome completeness is similar between NP swabs and saliva samples

To establish whether we could generate whole SARS-CoV-2 genomes from saliva, we used the ARTIC Network amplicon generation approach to sequence SARS-CoV-2 RNA extracted from 172 nasopharyngeal (NP) swabs and 60 saliva samples on the Oxford Nanopore MinION. We found that the cycle threshold (Ct) value for the N1 target according to the Centers for Disease Control and Prevention (CDC) SARS-CoV-2 RT-qPCR diagnostic assay is associated with genome completeness for both NP swabs and saliva samples (**Figure 1A**). Specifically, Ct values are inversely correlated with viral RNA quantity, and we could sequence more complete SARS-CoV-2 genomes from samples with low Ct values, regardless of sample type. We generated high-quality genomes from saliva samples with a wide range of Ct values. A Ct value of 30 is often used as a threshold by sequencing labs, where samples with higher Ct values may not generate complete SARS-CoV-2 genomes^11^. Based on our RT-qPCR standard curve, a Ct value of 30 corresponds to ∼1,000 SARS-CoV-2 genome equivalents (GE) per μL (**Methods**). Our data show that we could sequence the majority of samples with a Ct ≤30 to >80% completeness (165/172, 95.9%) (**Supplemental Table 1**). However, we generated fewer genomes with >80% completeness from saliva samples with a Ct ≤30 (35/41, 85.4%) compared to NP swabs (130/131, 99.2%).

**Figure 1.**
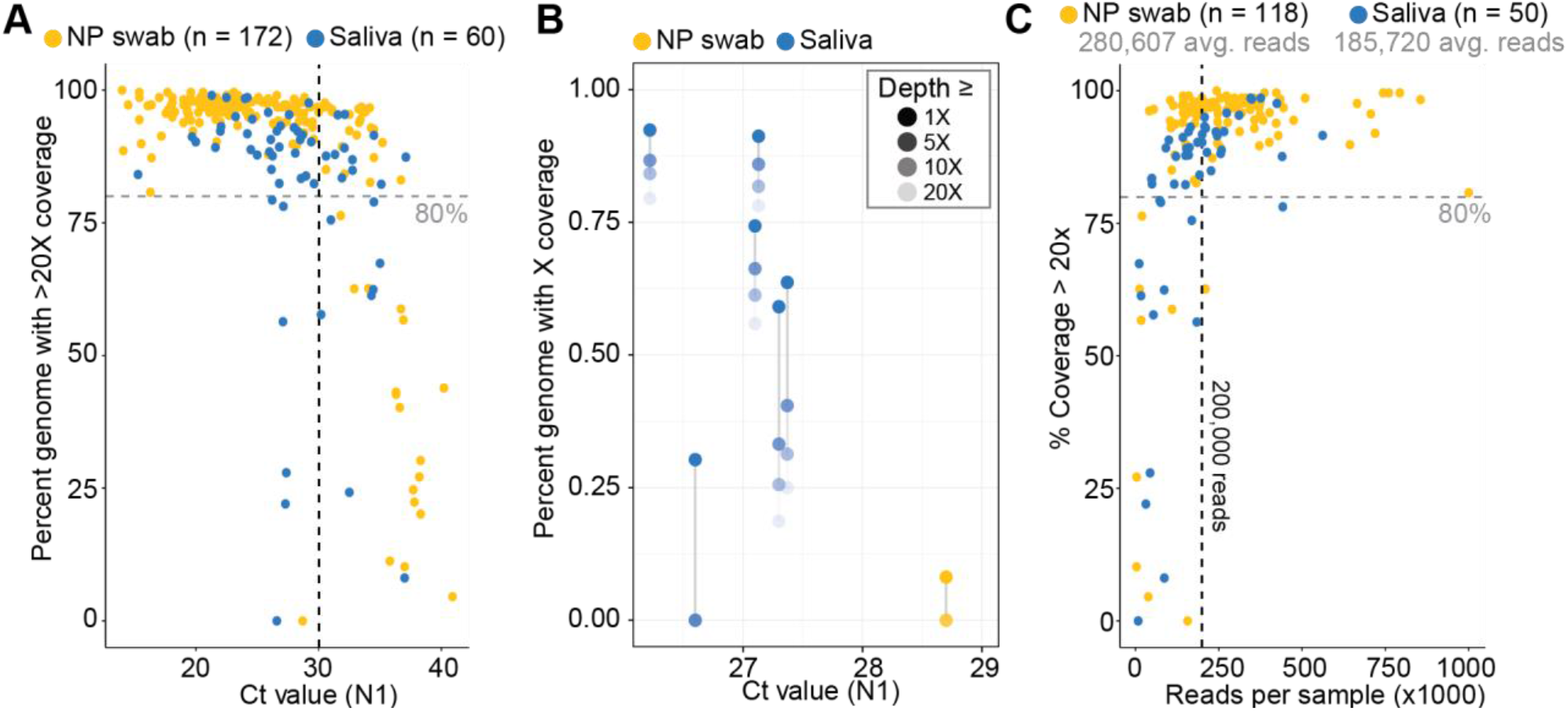
Saliva performs comparably to nasopharyngeal (NP) swabs as an original sample for SARS-CoV-2 genome sequencing. **A**. The percent of genome with at least 20x coverage is plotted against the Ct value for the N1 target for a cohort of unpaired saliva (blue) and NP swab (yellow) samples. Samples with a Ct value ≤30 (vertical black dashed line) and a genome completeness <80% (horizontal grey line) are displayed in panel B. **B**. The percent of the genome at different coverage thresholds (legend, top right) is plotted against Ct value for the N1 target for select samples from A. Grey lines connect points related to the same sample. **C**. A subset of samples from the cohorts in A are plotted against the number of reads for each sample, showing that nearly all samples (saliva and NP swab) with at least 200,000 reads (vertical black line) have >80% genome coverage. The mean readcount for each cohort is displayed underneath the legend.

As we require ≥20 aligned sequencing reads to call a base at any genome position, it is useful to assess lower depths of coverage to determine if more complete genomes would be generated with more data. We evaluated the discrepancy in SARS-CoV-2 genome completeness between sample types by calculating genome completeness at various depths of coverage for the 6 saliva samples and 1 NP swab with a low Ct value (≤30) and low genome completeness (<80%) (**Figure 1B**). For the 6 saliva samples at 10X, 5X, or 1X depth of coverage, we found that sequencing reads were generated across the genome, suggesting that simply allocating more sequencing space per sample would have resulted in more complete genomes. However, genome completeness was only slightly improved at lower depths of coverage for the NP swab sample.

We sequenced all samples included in this study in a multiplexed fashion, with as many as 22 samples included per run, and therefore the total amount of sequencing space given to each sample varies. As expected, we found a direct relationship between the total number of reads per sample and the genome completeness (**Figure 1C**). Of the 96 samples composed of > 200,000 reads, 94 (97.9%) exhibited high levels of genome completeness (>80%). Therefore we suggest allocating at least 200,000 reads per sample, regardless of sample type, to generate near-complete SARS-CoV-2 genomes using this sequencing approach.

### Sequencing matched saliva and NP swab samples from the same individuals produce genomes of similar completeness and relatedness

Next, we examined if saliva samples are directly comparable to NP swabs by sequencing matched samples taken from the same individual at the same time point. We obtained matched samples from 12 individuals and randomly downsampled the sequencing reads to match the sample with the least amount of data. We found no substantial difference in genome completeness for different sample types collected from the same individual (**Figure 2A**). For 5 individuals, saliva produced the more complete genome with normalized read counts compared to 7 for NP swabs. Saliva samples have a mean genome completeness of 91.1% ± 0.04% compared to NP samples at 92.0% ± 0.07%. Consistent with our previous results^6^, we found lower SARS-CoV-2 Ct values from saliva compared to the matched NP swabs. We also compared the genetic relatedness of every matched sample by constructing a maximum-likelihood phylogenetic tree. We found that 10 of the 12 pairs had identical genomes, where in two pairs (04 and 09) the genome sequenced from the NP swab had an additional mutation compared to the genome from saliva (**Figure 2B**). The mutations identified in the NP swab samples from pairs 04 and 09 are well supported with >400X depth of coverage at the sites. These mutations did not change the lineage placement for the NP swab genomes from samples 04-NP and 09-NP, both clustering with the B.1 pangolin lineage.

**Figure 2.**
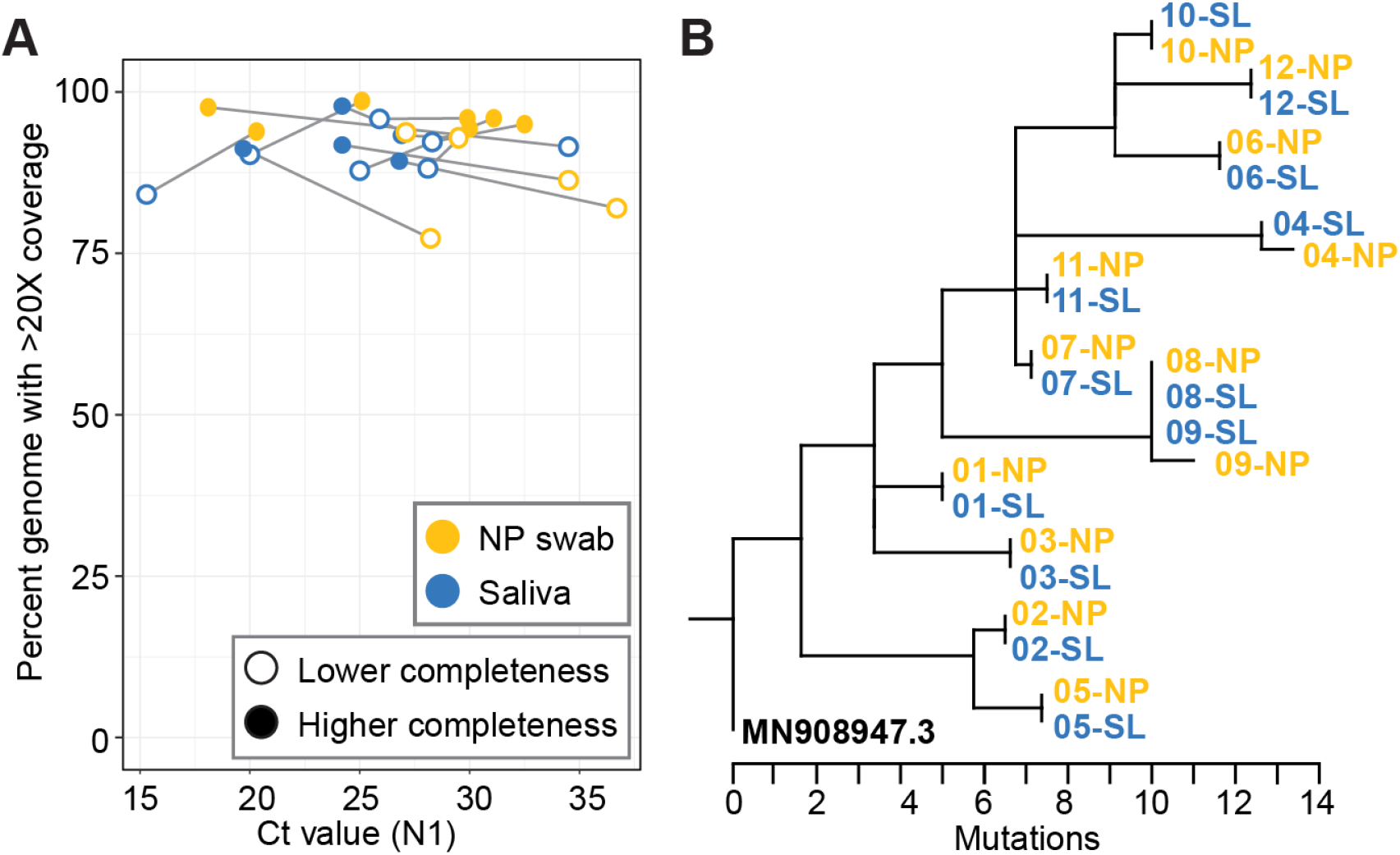
SARS-CoV-2 genomes from matched saliva and NP swabs are similar in completeness and content. **A**. A cohort of matched saliva and NP swab samples from the same individual were sequenced and reads were subsampled to match the mate with fewer reads. A grey line connects the mates and an empty circle highlights the mate with lower coverage. **B**. A maximum-likelihood tree of matched saliva and NP swab samples from Figure 2A is rooted against the reference genome (NCBI Accession MN908947.3) to show pairwise identity.

### RNA extraction improves completeness of SARS-CoV-2 genomes from saliva

The removal of the RNA extraction step for SARS-CoV-2 diagnostic test can streamline workflows and has been demonstrated for multiple sample types, including saliva^10,12,13^. SARS-CoV-2 genomic sequencing workflows would similarly benefit in the time and cost reductions by removing RNA extraction steps. We attempted to sequence SARS-CoV-2 genomes from split saliva samples, where half of the sample underwent our normal RNA extraction procedure and the other half was processed according to our SalivaDirect protocol with a proteinase K and heat treatment instead of RNA extraction^10^. While the extraction-free SalivaDirect protocol yielded similar SARS-CoV-2 Ct values to samples with RNA extraction, indicating similar levels of PCR detection, we found that sequencing from the SalivaDirect substrate led to a substantially lower genome coverage of 7.8% ± 33.3% compared to RNA extracted samples averaging 89.3% ± 29.0% completeness (**Figure 3A**). In fact, of the 11 samples processed using our SalivaDirect protocol, only 1 sequenced genome met our standards of 80% completeness, while the majority produced genomes that were less than 50% complete. In comparison to the counterpart sample that did undergo RNA extraction, we sequenced 9 out of 11 to >80% completeness, including some samples with Ct values >30. We then investigated whether this observation could be attributed to insufficient data by calculating the percentage completeness of each genome at various levels of depth of coverage (**Figure 3B**). Our analysis showed that increasing data per sample would improve genome coverage for only 4 of the samples processed with the extraction-free SalivaDirect protocol as evidenced by the large separation between data points in **Figure 3B**. However, 8 of these samples had no reads aligning to >90% of the genome, indicating that more data would not have substantially improved coverage, even at low Ct values. Thus, without further protocol optimization, RNA extract from saliva samples is necessary for sequencing complete SARS-CoV-2 genomes.

**Figure 3.**
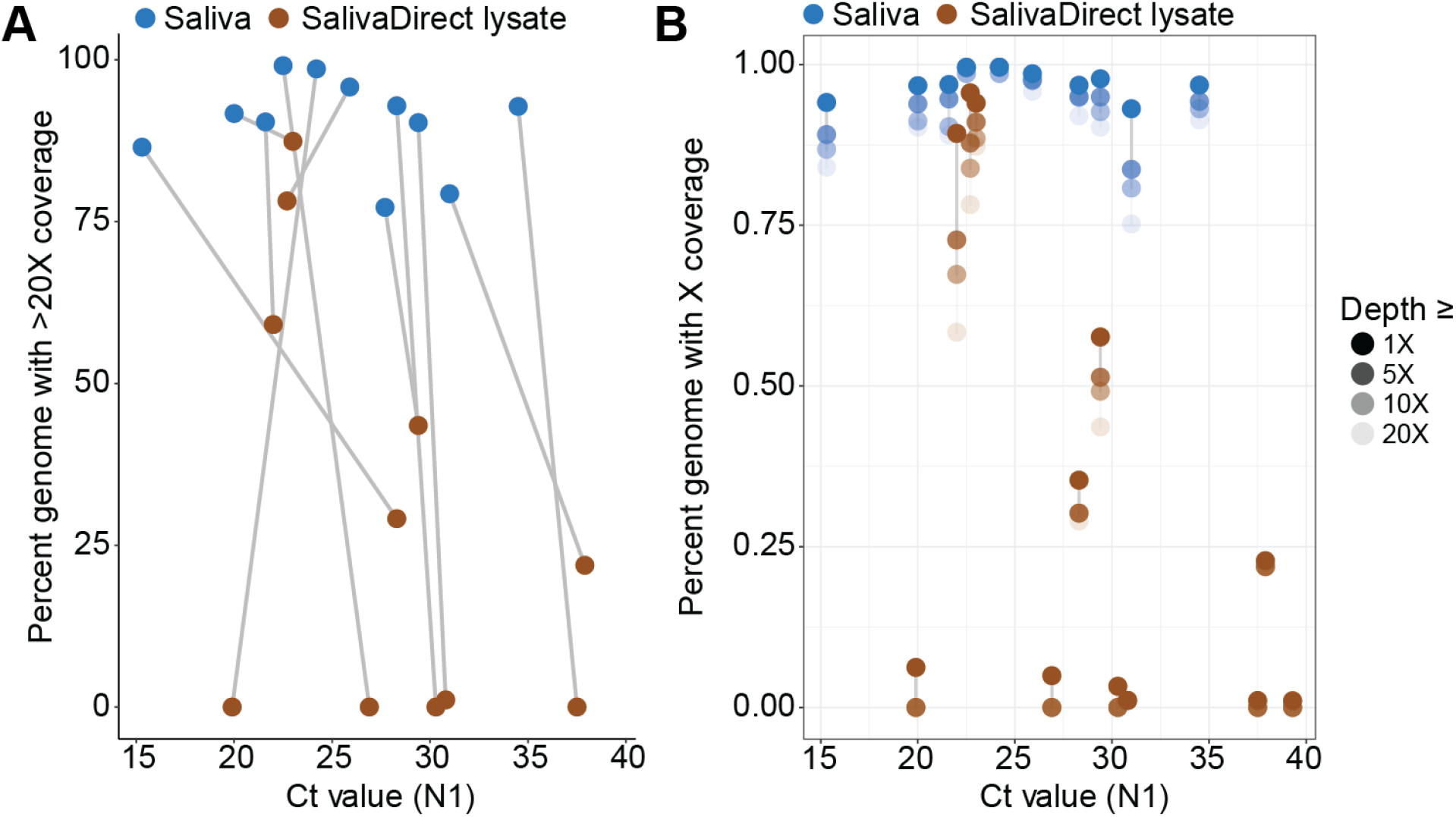
RNA extraction dramatically improves SARS-CoV-2 genome coverage from saliva samples. **A**. Saliva samples were split to perform either RNA extraction (blue) or SalivaDirect lysate (brown) preparation (incubation with Proteinase K at 95°C for 5 min; see methods) and were sequenced. The percent of genome with at least 20x coverage is plotted against the Ct value for the N1 target for matched samples (connected by grey line). **B**. The percent of the genome at different coverage thresholds (legend, right) is plotted against Ct value for the N1 target for the same cohort of samples in A. Grey lines connect points related to the same sample.

## Discussion

In this study, we determined that SARS-CoV-2 genomes generated from saliva were of the same completeness as those generated from NP swabs, the gold standard diagnostic specimen for COVID-19. By comparing a random subset of samples from saliva and NP swabs spanning different viral titers (**Figure 1A**), we found little difference between genome completeness from different sample types. Rather, viral titer (Ct values) and the amount of sequencing data generated from each sample is more indicative of genome completeness than specimen type (**Figure 1B**,**C**). Our results demonstrate that samples with a Ct value at or below 30, or ∼1000 SARS-CoV-2 GE/ul, with greater than 200,000 reads should produce near-complete SARS-CoV-2 genomes using using the ARTIC amplicon-based sequencing approach, in line with previously published data^11^. We observed substantial variation in Ct values from matched saliva and NP samples (**Figure 2A**), however the genome sequence itself was identical or nearly identical regardless of sample type (**Figure 2B**). When comparing saliva samples that have undergone RNA extraction to identical saliva samples processed with our SalivaDirect approach that excludes RNA extraction, we see a substantial drop in genome completeness (**Figure 3A**,**B**). This result may be explained by the highly multiplexed nature of the amplicon generation step being less efficient when RNA is not extracted and purified.

There are important considerations for the interpretation of these data. Our Ct value threshold of 30 is conservative and should not be seen as a lower limit of detection. Ct values will vary based on RT-qPCR assays, reagents, and thermocyclers, and we were able to sequence some near-complete SARS-CoV-2 genomes at Ct values greater than 30. Similarly, individual laboratory validation is necessary to determine the amount of data needed per sample if using protocols outside of the specific library preparation and sequencing approaches used in this study. Modifications of the primer concentrations used in the ARTIC Network’s amplicon-based sequencing approach have resulted in improved read distribution across the SARS-CoV-2 genome^4^. Our data were generated prior to these modifications, thus we would expect substantially fewer reads per sample will be needed to generate high-quality genomes. As well, data presented here was generated on the Oxford Nanopore MinION where the entire ∼400 b.p. amplicon is sequenced in a single read. Most library preparation approaches used for Illumina sequencing platforms fragment amplicons for shorter library insert lengths, meaning more reads will need to be generated per sample on short-read sequencing platforms.

In addition to these considerations, there are some limitations to our results. We did not assess how saliva performs as a specimen for sequencing SARS-CoV-2 genomes outside of amplicon-based strategies and additional research is needed to assess how saliva would perform in metagenomic or hybrid-/capture-based sequencing approaches. Additionally, we did not quantify the effect RNA extraction has on generating complete genomes from NP swabs. Therefore, we cannot determine if the reduction in genome completeness seen from samples without RNA is a specimen type specific phenomenon.

Saliva is a stable and sensitive specimen type for SARS-CoV-2 diagnostic assays. In addition to the safety afforded through self-collection opposed to NP swabs, saliva is increasingly becoming an appealing specimen type, in particular for large-scale public health surveillance. Taken together, our data indicate that saliva is a satisfactory sample type for not only diagnostic assays but also for sequencing high-quality SARS-CoV-2 genomes. This is an important finding for the scalability and streamlining of genomic epidemiological studies.

## Conclusions

The results of our study comparing the use of saliva to NP swabs for SARS-CoV-2 genome sequencing indicate that viral titers and data quantity influence genome completeness more than specimen type.

## Data Availability

All consensus genomes used in this study have been deposited at https://github.com/josephfauver/Saliva_Sequencing_Manuscript and are publicly available as of the date of publication. Accession numbers are listed in the key resources table.
All original code has been deposited at https://github.com/josephfauver/Saliva_Sequencing_Manuscript and is publicly available as of the date of publication.

## Yale IMPACT Research Team Members

Kelly Anastasio, Michael H. Askenase, Maria Batsu, Santos Bermejo, Sean Bickerton, Anderson Brito, Kristina Brower, Molly L. Bucklin, Staci Cahill, Melissa Campbell, Yiyun Cao, Arnau Casanovas-Massana, Edward Courchaine, Rupak Datta, Giuseppe DeIuliis, Charles Dela Cruz, Rebecca Earnest, Shelli Farhadian, Renata Filler, John Fournier, Bertie Geng, Laura Glick, Benjamin Goldman-Israelow, Ryan Handoko, Christina Harden, Akiko Iwasaki, Cole Jensen, Chaney Kalinich, William Khoury-Hanold, Daniel Kim, Jon Klein, Jonathan Klein, Lynda Knaggs, Albert Ko, Maxine Kuang, Eriko Kudo, Sarah Lapidus, Joseph Lim, Melissa Linehan, Feimei Liu, Peiwen Lu, Alice Lu-Culligan, Carolina Lucas, Amyn A. Malik, Tianyang Mao, Anjelica Martin, Irene Matos, David McDonald, Maksym Minasyan, Adam J. Moore, M. Catherine Muenker, Maura Nakahata, Nida Naushad, Allison Nelson, Jessica Nouws, Angela Nunez, Marcella Nunez-Smith, Abeer Obaid, Camila Odio, Ji Eun Oh, Saad Omer, Isabel M. Ott, Annsea Park, Hong-Jai Park, Xiaohua Peng, Sarah Prophet, Harold Rahming, Tyler Rice, Aaron Ring, Kadi-Ann Rose, Lorenzo Sewanan, Lokesh Sharma, Albert Shaw, Denise Shepard, Erin Silva, Julio Silva, Mikhail Smolgovsky, Eric Song, Nicole Sonnert, Yvette Strong, Codruta Todeasa, Maria Tokuyama, Jordan Valdez, Sofia Velazquez, Arvind Venkataraman, Pavithra Vijayakumar, Eric Y. Wang, Anne E. Watkins, Elizabeth B. White, Patrick Wong, Yexin Yang

## Acknowledgements

This work was funded by CTSA Grant Number TL1 TR001864 (T.A.), a fast grant from Emergent Ventures at the Mercatus Center at George Mason University (N.D.G.), and Centers for Disease Control and Prevention (CDC) contracts 75D30120C09570 (N.D.G.).

## Author Contributions

Conceptualization - T.A., C.B.F.V., A.L.W., N.D.G, J.R.F.; Investigation - T.A., C.B.F.V., M.I.B., M.E.P., J.R.F.; Writing - Original Draft - T.A., J.R.F.; Writing - Review & Editing - T.A., C.B.F.V., M.E.P., A.L.W., N.D.G., J.R.F.

## Methods

### RESOURCE AVAILABILITY

#### Lead contact

Further information and requests for resources and reagents should be directed to and will be fulfilled by the lead contact, Joseph R. Fauver.

#### Materials availability

This study did not generate new unique reagents.

#### Data and code availability

All consensus genomes used in this study have been deposited at https://github.com/josephfauver/Saliva_Sequencing_Manuscript and are publicly available as of the date of publication. Accession numbers are listed in the key resources table.

All original code has been deposited at https://github.com/josephfauver/Saliva_Sequencing_Manuscript and is publicly available as of the date of publication.

Any additional information required to reanalyze the data reported in this paper is available from the lead contact upon request.

### EXPERIMENTAL MODEL AND SUBJECT DETAILS

Nasopharyngeal swabs and saliva were collected from enrolled COVID-19 inpatients and healthcare workers from Yale-New Haven Hospital in accordance with the Yale University HIC-approved protocol #2000027690. Samples were collected after the study participant had acknowledged that they had understood the study protocol and signed the informed consent. All participant information and samples were collected in association with non-individually identifiable study identifiers. These samples were used to create the Yale COVID-19 Biorepository.

### METHOD DETAILS

#### Sample selection and RNA extraction

Samples were received as either purified RNA, original nasal swab in viral transport media, or original saliva sample. NP swab, saliva, and RNA samples came from the Yale-New Haven Hospital in partnership with the Yale Pathology Lab, Yale Clinical Virology Lab, and/or the Yale COVID-19 Biorepository. Nucleic acid was extracted from original samples (300 μL) using the MagMAX viral/pathogen nucleic acid isolation kit (Thermo Fisher) and eluted into 75 μL. Samples were screened for SARS-CoV-2 RNA via a multiplex RT-qPCR assay using the NEB Luna universal probe 1-Step RT-qPCR kit with CDC N1, N2, and RP primer-probe sets on the Bio-Rad CFX96 touch real-time PCR detection system^14^. PCR conditions were as follows: 55°C for 10 minutes, 95°C for 1 minute, 45 cycles of 95°C for 10 seconds followed by 55°C for 30 seconds. A standard curve of synthetic RNA transcripts was used to convert Ct values to genome equivalents (GE) (available on https://github.com/josephfauver/Saliva_Sequencing_Manuscript). Multiple extraction controls were included for each RNA extraction batch and tested negative for SARS-CoV-2 RNA by the same assay.

#### SalivaDirect

A detailed SalivaDirect protocol has been published^15^. Briefly, 50 μL of saliva is combined with 2.5 μL (50mg/mL) Proteinase K (Thermo Fisher) and vortexed for 1 min at 3200 RPM. After 5 minutes of incubation at 95°C for 5 minutes, 5 μL of lysate was directly used as input in the RT-qPCR reaction with the same conditions as specific for extracted RNA above. Samples were stored at -80°C before sequencing.

#### Oxford Nanopore library preparation and sequencing

RNA extracted from positive samples or processed SalivaDirect samples served as the inputs for an amplicon-based approach for sequencing on the Oxford Nanopore Technologies (ONT; Oxford, United Kingdom) MinION^16^. Sequencing libraries were prepared using the ONT Ligation Sequencing Kit (SQK-LSK109) and the ONT Native Barcoding Expansion pack as described in the ARTIC Network’s protocol with V3 primers (IDT)^17^ with the following modifications: cDNA was generated with SuperScriptIV VILO Master Mix (Thermo Fisher Scientific, Waltham, MA, USA), all amplicons were generated using 35 cycles of amplification, amplicons were then normalized to 15 ng for each sample, end repair incubation time was increased to 25 min followed by an additional bead-based clean up, and all clean up steps used a ratio of 1:1 beads:sample. No-template controls were introduced for each run at the cDNA synthesis and amplicon synthesis steps and were taken through the entire library preparation and sequencing protocol to detect any cross-contamination. For each control in each run, less than 1,000 total reads were observed. A subset of reads in control samples aligned to the SARS-CoV-2 genome, although no position of the genome had greater than 20 reads i.e. enough data to influence the generation of a consensus genome. 25 ng of the final library was loaded on a MinION R9.4.1 flow cell and sequenced for approximately 8-10 hours.

#### Bioinformatics processing

The RAMPART application from the ARTIC Network was used to monitor approximate genome coverage for each sample and control in real time during the sequencing run (github.com/artic-network/rampart). Fast5 files were basecalled using the Guppy basecaller 4.4.0 fast model and consensus genomes were generated according to the ARTIC bioinformatic pipeline (artic.network/ncov-2019/ncov2019-bioinformatics-sop.html) which uses Nanopolish to call variants^18^. A threshold of 20x coverage was required to call a base pair in the consensus genome. Regions of the genome with less than 20x coverage were designated with N’s in the consensus genome.

### QUANTIFICATION AND STATISTICAL ANALYSIS

Genome completeness was determined either by counting N’s in the consensus genome output by the ARTIC pipeline for 20x coverage (Figure 1A and C) or by using the SAMtools^19^ depth feature for various coverage thresholds (Figure 1B). At the 20x threshold, these two methods produced highly similar values with a slight discrepancy.

Matched saliva and NP swab genomes were aligned with MAFFT^20,21^ and placed in a phylogenetic tree using a maximum-likelihood approach (PHYML) to show similarity between matched samples^22^. Some manual curation was necessary to correct for misalignment over ambiguous regions of the genome. Lineage assignments were conducted using pangolin (v.3.1.3)^23^

## Tables with titles and legends

**Supplementary Table 1:**
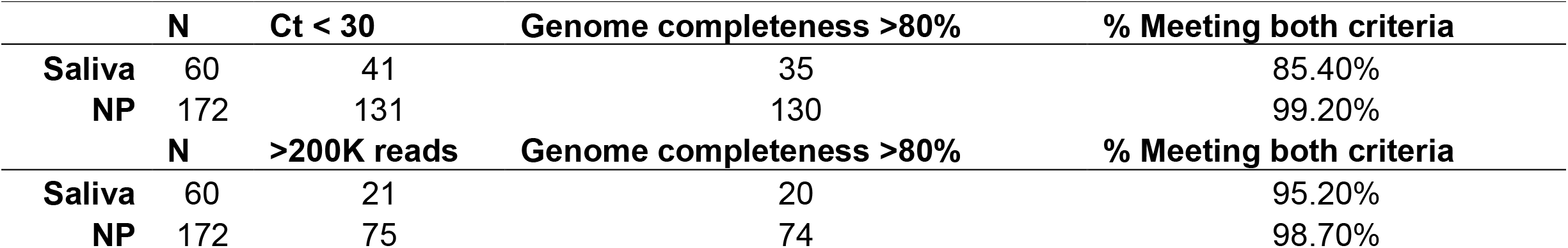
Summary statistics from saliva or nasopharyngeal (NP) swab samples.

